# Design and evaluation of probe-capture sequencing for clinical *Neisseria gonorrhoeae* specimens

**DOI:** 10.64898/2026.10.27.26366443

**Authors:** Aryan Singh, Kyle J. Gontjes, Dhatri Badri Narayanan, Paul M. Won, Brian W. Brunelle, Stefan J. Green, Kyle J. Popovich, Evan S. Snitkin

## Abstract

Characterizing the molecular epidemiology of *Neisseria gonorrhoeae* is essential for understanding disease burden, elucidating transmission dynamics, and informing clinical decision-making and public health interventions. Given that routine, culture-independent diagnostics rapidly and sensitively detect this difficult-to-cultivate pathogen, aligning genomic surveillance with these workflows is necessary to understand its epidemiology and evolutionary dynamics. To address this challenge, we investigated the utility of probe-capture sequencing to selectively enrich for pathogen-specific DNA during next-generation sequencing. Custom biotinylated oligonucleotide baits were designed using genetic information from publicly available genomes hosted on Pathogenwatch. Of the 32,574 assemblies, 2,877 representative genomes were selected to inform probe generation using core genome amino acid distance clustering and representation of unique resistance-associated genotypes and predicted resistance phenotypes. The resulting probe set provided generous coverage of the reference genome, *N. gonorrhoeae* FA 1090, with 94.6% nucleotide coverage and 99.3% of annotated genes covered by at least one probe. Piloting this technology on 12 clinical urine specimens with variable relative pathogen abundances revealed that probe-capture sequencing substantially increased the relative abundance of *N. gonorrhoeae* sequence reads from a complex sample (fold-change range, 226 - 2046), enabling considerable genome coverage, the construction of high-quality genome assemblies, and the detection of resistance-associated genotypes. These findings demonstrate that probe-capture sequencing is a viable method for enriching *N. gonorrhoeae* genetic material from clinical samples, enabling culture-independent genomic investigations into the epidemiology of hard-to-culture pathogens and providing actionable insights to inform clinical and public health interventions.

**IMPORTANCE:** Understanding the molecular epidemiology of bacterial pathogens can illuminate population diversity, transmission dynamics, and genotypes associated with bacterial traits and clinical outcomes. Difficulties in cultivation hinder our ability to understand the epidemiology of *Neisseria gonorrhoeae*. Aligning genomic surveillance methodologies with existing clinical diagnostic modalities is essential to guiding the implementation of innovative genomic surveillance. In this paper, we used representative public genome assemblies to design custom biotinylated oligonucleotide baits to successfully increase pathogen-specific DNA yield from complex clinical urine specimens during next-generation sequencing. Probe-capture sequencing markedly improved detection of pathogen-specific genetic material from clinical specimens, enabling construction of high-quality genome assemblies and detection of resistance-associated genotypes. Collectively, we demonstrate that probe-capture sequencing is a viable method for sequencing hard-to-culture pathogens from clinical specimens, highlighting how this technology could strengthen genomic investigations into the epidemiology of these pathogens and provide actionable insights to inform clinical decision-making and public health interventions.

## INTRODUCTION

*Neisseria gonorrhoeae* is a Gram-negative diplococcus that infects exposed mucosal surfaces. As the causative agent of gonorrhea, a sexually transmitted infection, this pathogen is of global public health significance, with the World Health Organization estimating 82 million new infections among adults in 2020 (1, 2). As *N. gonorrhoeae* can evolve resistance to all clinically relevant antibiotics (3, 4), the emergence and spread of multidrug-resistant lineages, especially those resistant to extended-spectrum cephalosporins, emphasizes the need for a sharpened understanding of its epidemiology and evolution (5).

*N. gonorrhoeae* is challenging to cultivate, and this has limited full exploration of the epidemiology of this pathogen and similarly fastidious microorganisms (6). To facilitate the rapid and accurate detection of *N. gonorrhoeae*, laboratories use nucleic acid amplification tests (NAAT), the standard diagnostic for infection in the United States (6). Although NAAT permits rapid and sensitive detection, routine diagnostic testing alone provides limited information on genetic diversity and antibiotic resistance. Culture-dependent surveillance can ameliorate this concern; however, reliance on culturing biases downstream analyses toward anatomical sites and lineages with higher recovery rates under laboratory conditions (7). Metagenomic sequencing represents another promising approach for pathogen detection; however, low bacterial abundance relative to host DNA and commensal microbiota can complicate the recovery of sufficient pathogen-specific reads for reliable surveillance (8). As a result, genomic epidemiology must adapt to support surveillance efforts while preserving the genomic content needed to analyze community trends in a timely manner and guide public health action.

Probe-capture sequencing can circumvent these limitations. The design of biotinylated oligonucleotide baits to selectively enrich target DNA prior to sequencing can increase read recovery from complex, host-dominated specimens (9). Similar hybridization-based approaches have been successfully applied to microbial and non-microbial sources, demonstrating the utility of targeted enrichment for improving sequencing depth and genome recovery from challenging sample types (9, 10). For clinical and public health microbiologists, this technology can be leveraged to identify rare antimicrobial resistance determinants, strengthen differential diagnosis, and reveal disease-associated responses of low-abundance bacteria within their native environments (10-12).

Here, we pilot a probe-capture sequencing approach for the detection of *N. gonorrhoeae*. To enhance pathogen capture, we generated a custom probe set based on genome sequences from 2,877 representative, publicly available genomes. We piloted this technology on twelve clinical urine specimens with varying relative abundances of *N. gonorrhoeae*. This study shows that targeted enrichment improves pathogen recovery from complex clinical specimens, offering an informative genomic surveillance approach for this hard-to-culture pathogen.

## METHODS

### Selection of representative *Neisseria gonorrhoeae* genomes for probe design

As probe-capture sequencing performance declines in sensitivity and depth of coverage in divergent populations (13), it is critical to generate a set of probes that accounts for genetic diversity. To inform the design of custom, biotinylated RNA probes for *N. gonorrhoeae*, we downloaded assemblies from 30 Pathogenwatch collections (https://pathogen.watch/) on November 12, 2024. Pathogenwatch was leveraged for its spatiotemporal diversity and uniform bioinformatic pipelines, including in-house genome annotation, multi-locus sequence typing, antimicrobial resistance genotyping, and predicted resistance phenotypes (14). Continent and country data were extrapolated from location metadata using rnaturalearth v1.2.0 (15).

To identify representative genomes to inform probe design, genomes were clustered using core gene amino acid distance (**Figure 1A**). Core gene annotation and alignment were performed using cognac v.1.0 (16). An amino acid sequence alignment of 1,426 core genes, defined as genes present in ≥ 95% of genomes, was generated from 32,574 genomes. To visualize relationships among genomes, a neighbor-joining phylogeny was built from the amino acid distance matrix using ape (17). Next, genomes were grouped into genetically related clusters, using complete-linkage clustering with an amino acid distance threshold of 350, chosen after examining how cluster number and size varied across thresholds (**Supplementary Figure 1)**. To account for intra-cluster diversity, multi-member groups were further partitioned based on detected antimicrobial resistance-associated genotypes and predicted resistance phenotypes. Resistance-associated genotypes were evaluated as the presence of any mutation in the locus. An example of amino acid and trait-based partitioning from this collection is provided in **Figure 1B**.

**Figure 1.**
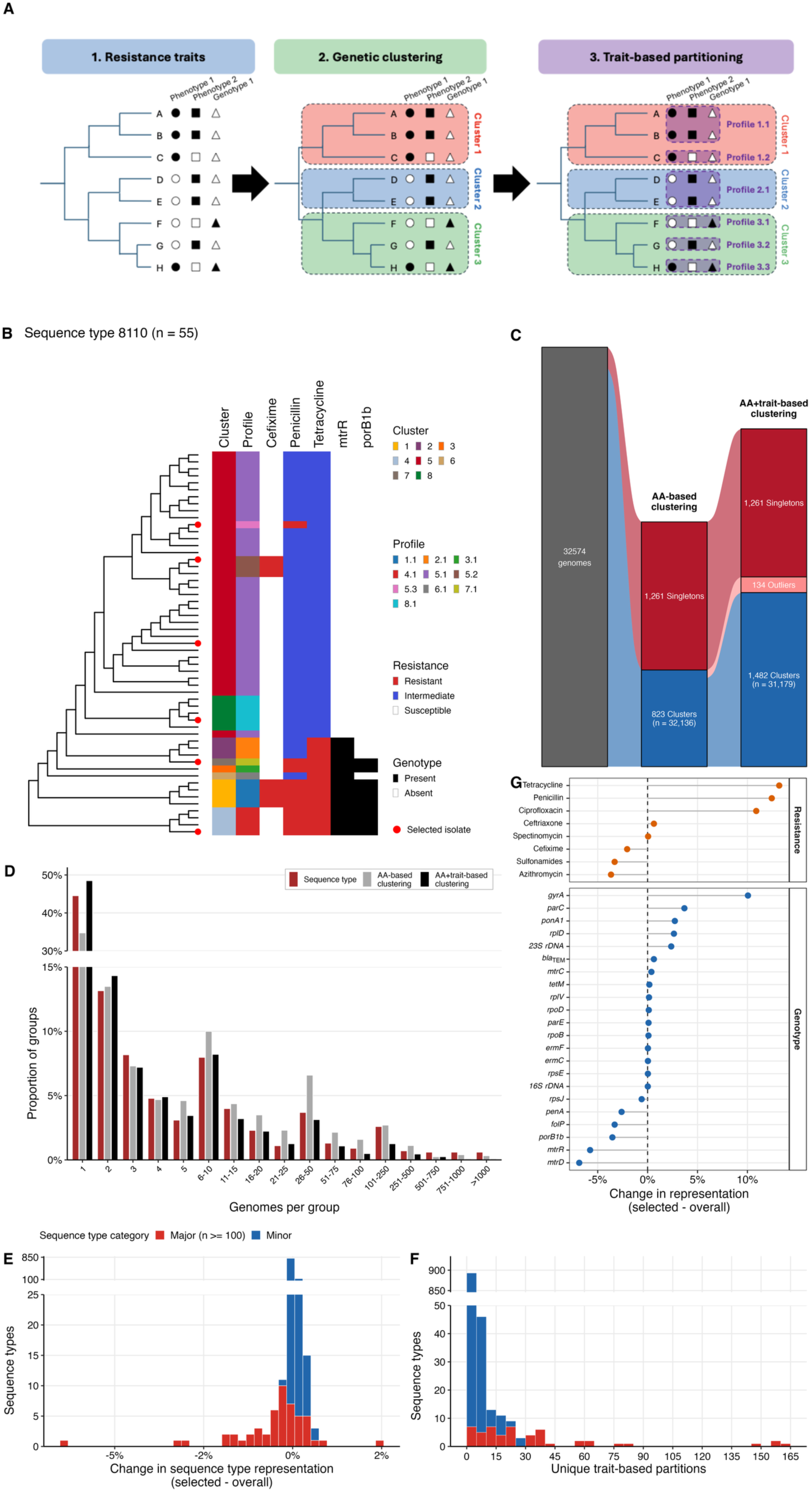
Genomic diversity and representativeness of selected isolate collection. (A) Methodology used to partition isolates into informative groups. Genomes are partitioned into clusters using a distance threshold. Next, isolates belonging to the same amino acid (AA) distance-based cluster are partitioned into groups based on the relevant traits, such as resistance-associated genotypes and phenotypes. This methodology was implemented using available *Neisseria gonorrhoeae* genomes downloaded from Pathogenwatch (November 12, 2024). Core-gene alignment of 32,574 genomes was performed, with an amino acid distance threshold of 350 used to partition isolates into amino acid-based clusters. Resistance-associated genotypes and phenotype patterns were used to partition cluster isolates into trait-based groups. (B) Genome partitioning among a representative *N. gonorrhoeae* sequence type. (C) Alluvial plot of the assignment of study genomes into amino acid-based clusters (AA-based clustering) and further partitioning using resistance-associated genotype and phenotype patterns (AA+trait-based clustering). Clusters were defined as groups with more than one genome. (D) Distribution of group sizes among three partitions: sequence type, amino acid distance-based clustering, and further partitioning by resistance-associated traits. (E) Change in sequence type representation when comparing the selected genomes and the overall collection. Major sequence types were defined as sequence types representing more than 1% of the population. (F) Number of unique trait-based partitions among sequence types. (G) Difference in the prevalence of resistance phenotypes and resistance-associated genotypes between selected and overall genome collections, expressed in percentage points.

### Design and synthesis of custom hybridization capture sequencing probes for *Neisseria gonorrhoeae*

After curating genetically related *N. gonorrhoeae* subpopulations with shared antibiotic resistance patterns, we used assembly quality control data to select high-quality genomes to guide probe generation. For each multi-assembly group, the assembly with the fewest contigs and the highest N50 was selected. A total of 2,877 genome assemblies were selected to inform the design of custom hybridization-capture sequencing probes using Daicel Arbor Biosciences’ myBaits Custom DNA-Seq platform (Daicel Arbor Biosciences, Ann Arbor, MI). To assess the representativeness of these genomes, spatiotemporal, sequence type, and antimicrobial resistance metadata were compared between the selected genomes and the overall Pathogenwatch dataset using Fisher’s exact test.

Probes were designed using 120 nt sequences with 2x tiling (overlapping probes every ∼60 nt along each target sequence) from all 2,877 genome assemblies; probe positions shifted slightly where necessary to distribute probes evenly along each target locus. After keeping unique sequences, probe sequences were soft masked for low complexity and simple repeats (0.3% of all sequences were soft masked) using RepeatMasker v4.1.3-p1. To identify probes that may cross-hybridize with highly abundant exogenous DNA that is potentially present in the urine samples, blastn v2.12.0+ was used to align probe sequences to the following genomes: human, *Escherichia coli*, *Pseudomonas aeruginosa*, *Enterococcus faecalis*, *Ralstonia solanacearum*, *Haemophilus ducreyi*, *Staphylococcus haemolyticus*, *Aeromonas jandaei*, *Klebsiella variicola*, *Staphylococcus aureus*, *Klebsiella pneumoniae*, and *Bacillus paralicheniformis* (18). Only probes that did not align to any of the genomes and were ≤35% soft-masked for simple repeats were retained. To reduce the probe count without affecting performance, sequences were randomized, and an in-house greedy clustering algorithm was employed to group sequences that were > 96.5% identical over at least 75% of the sequence. One representative sequence per cluster was retained, as this sequence is expected to hybridize with comparable efficiency to all corresponding targets that are ≥ 95% identical.

To evaluate coverage of the custom probe set, all probe sequences were aligned to the well-characterized reference genome *N. gonorrhoeae* FA 1090 (GenBank accession NC_002946.2) using BWA-MEM (19). Alignments were converted into BED format using SAMtools v.1.19.2 and BEDTools v2.17.0 (20, 21). Genomic coverage was assessed at the nucleotide level (fraction of nucleotides overlapped by at least one probe) and gene level (proportion of annotated genes covered, and fraction of each gene’s coding sequence covered, by at least one probe).

### Application of probe-capture sequencing to clinical urine specimens

To pilot this methodology, probe-capture sequencing was performed on twelve *N. gonorrhoeae*-positive clinical urine culture specimens. DNA was extracted from urine specimens using a Maxwell RSC Cultured Cells DNA kit (Promega, Madison, WI) implemented on a Maxwell RSC device, according to the manufacturer’s instructions. Extracted genomic DNA (gDNA) was used as input for shotgun library preparation using an NEXTFLEX Rapid DNA-Seq Kit 2.0 kit and implemented on a Sciclone G3 NGSx iQ instrument (Revvity; Revvity, Waltham, MA). Libraries were amplified using an icon96 instrument (n6tec) with auto-normalization using the slope method. Libraries were initially sequenced shallowly (∼1 M clusters/samples) using a portion of an Illumina NovaSeq X lane (300 cycle chemistry) to evaluate approximate relative abundance of *N. gonorrhoeae* sequences prior to capture, with transcripts from the *N. gonorrhoeae* strain FDAARGOS_204 reference genome (GenBank accession CP020415) used as a rapid proxy. Based on these shallow metagenome sequence datasets, specimens were chosen to represent low-(<0.01%), medium-(0.01-0.1%), and high-(>0.1%) pathogen abundance. Libraries were pooled in batches of 4 for each abundance class in approximate equimolar ratios of 250 ng (range of 144-250 ng/sample) to a final mass of 846 to 957 ng total per capture. Each pool was polymerase chain reaction (PCR)-amplified with P5/P7 primers to increase total library amount, bead-purified with 0.8X bead volume (KAPA Pure beads, Roche) and eluted in 7 µl. Captures were performed using the myBaits Custom Hybridization Capture protocol according to the manufacturer’s instructions, including post-capture amplification. Post-capture libraries were deeply sequenced on an Illumina NovaSeq X instrument, using 300-cycle chemistry. Extraction, library preparation, and capture were performed in the Genomics and Microbiome Core Facility (GMCF; RRID:SCR_026856). NovaSeq X sequencing was performed at the DNA Services Core at the University of Illinois at Urbana-Champaign. The Design ID (DID) for this probe set (v1) is D10891NGNRH.

### Probe-capture sequencing genomic data processing

Post-capture raw paired-end reads underwent quality control, genome assembly, and annotation using NG-probe, a custom, in-house Snakemake pipeline (https://github.com/Snitkin-Lab-Umich/NG-probe) (22, 23). The depletion of host DNA was performed by KneadData v.0.12.3 using the hg39 reference database (24). PCR duplicates were removed from host-depleted reads using BBTools’ clumpify v.39.26 (25). Subsequently, taxonomic profiling was performed using Kraken2 v.2.1.6 (26). To confirm the removal of low-quality bases, adapters, and other contaminants, pre- and post-processing read quality was assessed using FastQC v.0.12.1 (27).

To extract pathogen-specific reads, filtered reads were mapped to an *N. gonorrhoeae* genome database comprising the 2,877 concatenated genomes used for probe design via Bowtie *2* v.2.5.4 (28). To calculate genome coverage, extracted reads were realigned to a single, randomly selected reference genome (SAMEA12279355) from the database, and the proportion of bases covered at various depth thresholds was calculated. Mapped reads were retained for downstream genome assembly and annotation. Specifically, mapped reads were downsampled using seqtk v1.0-r82-dirty (29). Genome assemblies were produced using *SPAdes* v.4.0.0 (30), with contigs shorter than 1 kb removed using bioawk v1.0 (31). Assembly quality and genome completeness were measured using QUAST v.5.3.0 and BUSCO v5.8.2, respectively (32, 33). Species identification and sequence typing were performed using skani v0.2.1 and mlst v2.23.0, respectively (34, 35).

To evaluate the detection of resistance genotypes and alleles, we performed antimicrobial resistance genotyping using Pathogenwatch’s AMRsearch v.1.1.4 (14). Next, read mapping to the *N. gonorrhoeae* AMRsearch database and *N. gonorrhoeae* FA 1090 reference genome was performed using the custom, inhouse Snakemake pipeline (https://github.com/kylegontjes/CoverageStats). To minimize cross-mapping, reads were mapped to each resistance-associated reference gene individually. Briefly, reads were mapped to the reference using bwa v.0.7.17 and converted to BAM files using SAMtools v.1.19 (20). GATK’s DepthOfCoverage tool was used on polymerase chain reaction duplicate-removed files to calculate coverage to the reference (36). For each resistance locus and documented Pathogenwatch resistance allele, chromosome-normalized median non-zero depth was calculated as the median non-zero depth across the region divided by the median non-zero depth across the *N. gonorrhoeae* FA 1090 reference genome.

### Data analysis and visualization

All analyses were performed in R v.4.6.1, unless otherwise noted. Data were cleaned and visualized using *tidyverse* packages and ggnewscale v0.5.2 (37, 38). Phylogenetic reconstruction, analysis, and visualization were performed using phytools v.2.5-2 and ape v.5.8-1 (17, 39). Multi-panel figures were generated using cowplot v.1.2.0 (40).

## RESULTS

### Characterization of a global collection of publicly available *Neisseria gonorrhoeae* genomes

To create a custom set of probes for capture sequencing, 32,574 *N. gonorrhoeae* genomes were downloaded from 30 Pathogenwatch collections (**Supplementary Figure 2**). Most genomes were collected between 2010 and 2020 (23,722; 80.8%), with a median collection year of 2017 (interquartile range [IQR], 2014-2019; missing collection date, 3,222). Among isolates with location metadata (29,149; 89.5%), the largest number were obtained from the Americas (14,014; 48.1%). Of the 68 represented countries, the five largest contributors were the United States (12,203; 37.5%), United Kingdom (3,702; 11.5%), Australia (2,418; 7.4%), Norway (2,295; 7.0%), and Sweden (1,273; 3.9%).

Multilocus sequence typing revealed 996 sequence types (STs), with the most abundant being ST9363 (3,734; 11.5%), ST1901 (2747; 8.4%), ST7363 (1540; 4.7%), ST8156 (1407; 4.3%), and ST8143 (1052; 3.2%). Antibiotic non-susceptibility predictions were common for penicillin (96.0%), tetracycline (89.3%), sulfonamides (86.1%), and ciprofloxacin (44.7%), while non-susceptibility was infrequent for azithromycin (14.8%), cefixime (8.7%), ceftriaxone (0.4%), and spectinomycin (0.02%). Sixty-two resistance-associated mutations spanning 22 loci were identified, with frequencies shown in **Supplemental Figure 2E**. Inspection of resistance-associated genotypes and predicted resistance patterns across genomes revealed 1,807 distinct patterns. After grouping resistance-associated mutations at the gene level, this number decreased to 592.

### Generation of a representative sample of publicly available isolates to inform probe design

To generate a representative sample of assemblies for probe-capture sequencing, 32,574 genomes were clustered into genetically similar groups based on core-gene amino acid distance. Unsurprisingly, as the amino acid threshold increased, cluster membership increased, with all assemblies belonging to a cluster at the distance threshold of 1,225 (**Supplementary Figure 1A)**. Cluster size also increased as the threshold increased, consistent with progressive grouping of distinctly related assemblies (**Supplementary Figure 1B**).

To curate a more representative grouping of assemblies, minimize redundancy, and maximize inclusion of resistance determinants in probe design, genetic clusters were partitioned based on resistance-associated genotype and predicted resistance patterns (**Figure 1)**. An amino acid distance threshold of 350 was employed to balance the retention of intra-cluster genetic diversity and limit excessive merging of distantly related isolates. At this threshold, 823 clusters comprising 32,136 genomes and 1,261 singletons were identified. Partitioning clusters by their resistance-associated genotype and predicted resistance patterns generated a total of 2,877 distinct groups (**Figure 1C)**. Resistance-associated genotype- and phenotype-informed grouping resulted in many more distinct groups of smaller sizes (**Figure 1D**), underscoring within-group variation in genotypic and phenotypic diversity and the need to use bacterial metadata when selecting isolates for probe design. The phylogenetic distribution of selected isolates and their resistance-associated genotypic and resistance predictions are shown in **Supplementary Figure 3**, with their placement within the full study tree highlighted in **Supplementary Figure 4.** Spatiotemporal, sequence type diversity, and resistance-associated traits of selected isolates are reported in **Supplementary Figure 5**.

### Chosen assemblies capture genotype distributions despite shifting lineage compositions

To evaluate whether the chosen assemblies reflected the diverse global collection, we compared the temporal, geographic, ST, and resistance patterns across the two datasets (**Supplementary Figure 6)**. Although collection decade distribution differed across collections (Fisher’s exact test p-value < 0.001), most were collected between 2010 and 2020 (chosen, 78.1%; overall, 80.8%). Changes in geographic representation were detected, notably increases in the proportion of isolates from Africa (chosen, 5.7%; overall, 1.1%) and Asia (chosen, 9.3%; overall, 3.5%) and decreases in representation of genomes from the Americas (chosen, 43.7%; overall, 48.1%) and Oceania (chosen, 4.8%; overall, 10.7%). Sixty-one percent (610/996) of STs detected in the global collection were present in the representative collection. Although the ST distribution significantly differed (Fisher’s exact test p-value <0.001), this was driven by increased representation of less abundant STs (**Figure 1E)**. Furthermore, some STs had greater diversity in resistance-associated genotype and phenotype patterns (**Figure 1F**). Non-susceptibility patterns differed between collections for most antibiotics (**Figure 1F & Supplementary Figure 6D**). Specifically, non-susceptibility to ceftriaxone and ciprofloxacin increased, while non-susceptibility to azithromycin and sulfonamide decreased (Fisher’s exact test p-values <0.001). Similarly, differences in the presence of resistance-associated genotypes were also detected (**Supplementary Figure 6E).** Collectively, major resistance patterns remained represented, but their distributions shifted in the collection, consistent with altered representation of less abundant genetic backgrounds (**Figure 1D-G)**.

### Custom probe set achieved comprehensive coverage of a well-characterized reference genome

Using the 2,877 representative genomes, 1,006,000,961 probes (120 nt every ∼60 nt along each genome) were designed. Exclusion of redundant sequences reduced the number to 5,539,042 probes. To remove probes that could saturate read data and obscure the target data, probes containing low complexity, sequence repeats, or aligned to other bacterial species likely to be prevalent in the sample were removed, resulting in 5,039,463 probes. After clustering probes into representative nodes based on sequence identity, 139,333 probe sequences were generated.

Probe set coverage was evaluated against the well-characterized *N. gonorrhoeae* FA 1090 reference genome (**Figure 2**). The probes directly covered 94.6% of the genome (2.04 / 2.15 Mbp), with 99.3% (2234 / 2250) of annotated genes covered by at least one probe; as probes enrich both the target and flanking regions, the effective coverage is even higher. Seventy-six percent (1708 / 2,250) of genes were fully covered by probes (**Figure 2A-B**). Partially covered genes (526) were predominantly covered at least 80% (340 [64.6%]), with the 5’ and 3’ ends having the best coverage across the genes (**Figure 2C-D**). Investigation of resistance-associated genotypes in the reference genome revealed that 9 had full coverage, with the remaining having numerous stretches of probe coverage, including start and stop regions (**Figure 2E**). However, very low coverage was detected across ribosomal RNA genes: 16S rRNA gene (range, 0.0%-5.2%) and 23S rRNA gene (range, 0.0%-9.1%).

**Figure 2.**
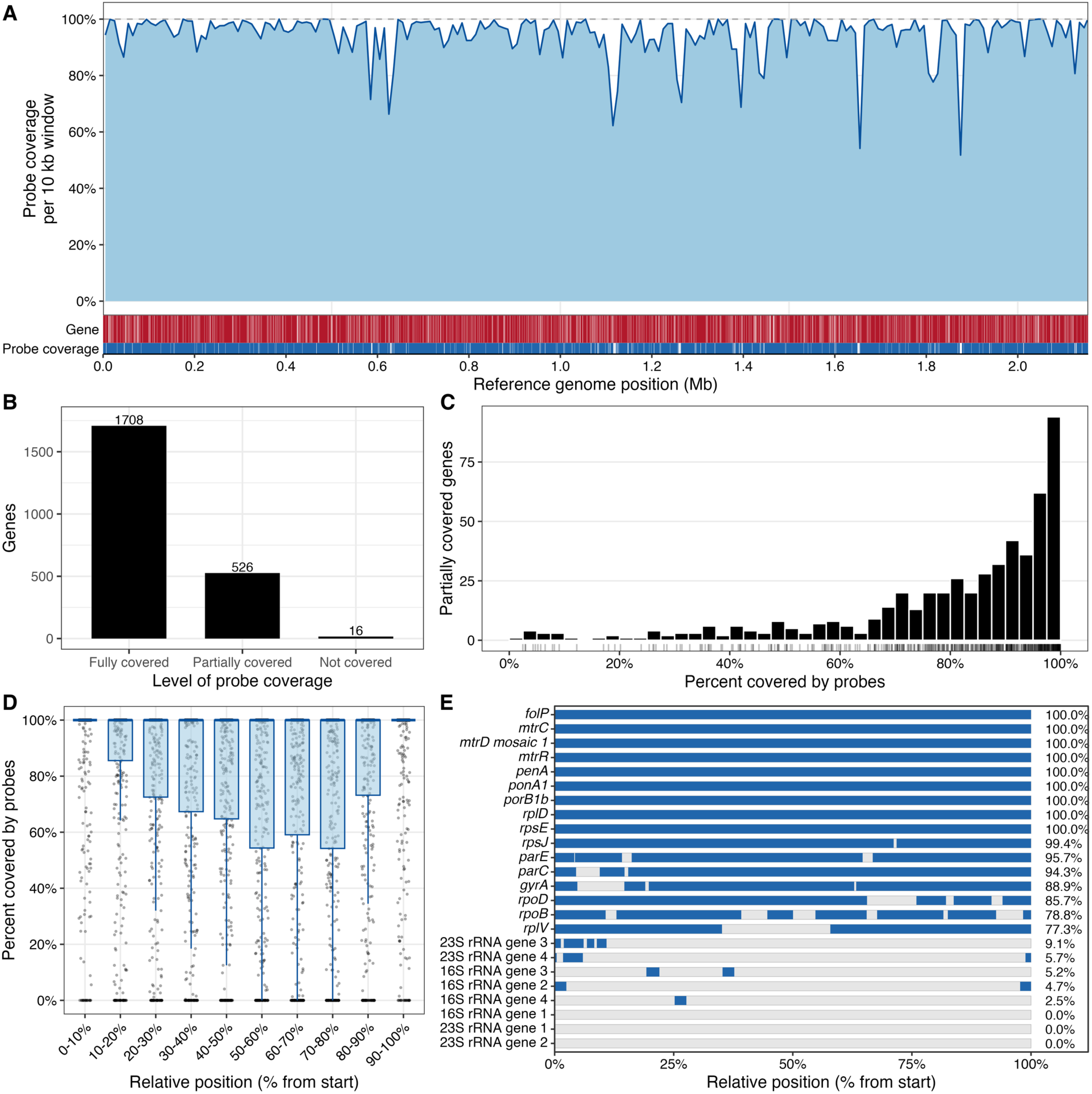
Coverage of the probe set across a representative *Neisseria gonorrhoeae* reference genome. Probe coverage was evaluated via read mapping against the *Neisseria gonorrhoeae* FA 1090 reference genome. (A) Percentage of the reference genome covered by probes in 10kb windows. Gene and probe heatmaps appended indicate the genomic positions of annotated genes and aligned probes, respectively. (B) Alluvial plot summarizing the probe coverage patterns of 2,250 annotated genes. (C) Distribution of the percent coverage of partially covered genes. Bars indicate the number of partially covered genes in the 2.5% coverage range, with ticks representing the percent covered for an individual gene. (D) Distribution of the probe coverage across the relative positions of the partially covered genes. Genes were divided into 10 equal-length bins from 5’ to 3’; points represent the individual gene and boxplots summarize their distributions per bin. (E) Probe coverage across the resistance-associated loci. Blue indicates probe-covered regions, light gray denotes uncovered regions, and values summarize the overall percentage of each locus covered.

### Probe-capture sequencing recovers *Neisseria gonorrhoeae* genetic material from clinical specimens

To evaluate genome recovery in clinical specimens, probe-capture sequencing was applied to 12 clinical urine specimens, stratified into low-, medium-, and high-abundance categories based on the relative abundance of *N. gonorrhoeae* in sample libraries prior to hybridization capture enrichment. Preliminary evaluation of probe-capture sequencing using shallow sequencing revealed considerable enrichment of pathogen-specific DNA (fold change, 226 – 2,046) (**Figure 3A**). Read counts markedly decreased following trimming and human-read removal, with higher-abundance specimens retaining more reads after subsequent pathogen-specific read retrieval (**Figure 3B)**. Despite differences in initial pathogen abundance, probe-capture sequencing substantially increased the relative pathogen abundance across most specimens, with a median post-enrichment relative abundance of 24.7% (IQR, 2.2%-50.7%) (**Figure 3C**). Genome coverage was high among read-depth thresholds for high- and medium-abundance specimens, while coverage declined more substantially in the low-abundance specimens (**Figure 3D**). Most specimens (9/12) produced assemblies approaching expected *N. gonorrhoeae* genome size (∼2.1-2.2 Mbp) (41), while low-abundance specimens generated smaller and more fragmented assemblies (**Figures 3E-F**).

**Figure 3.**
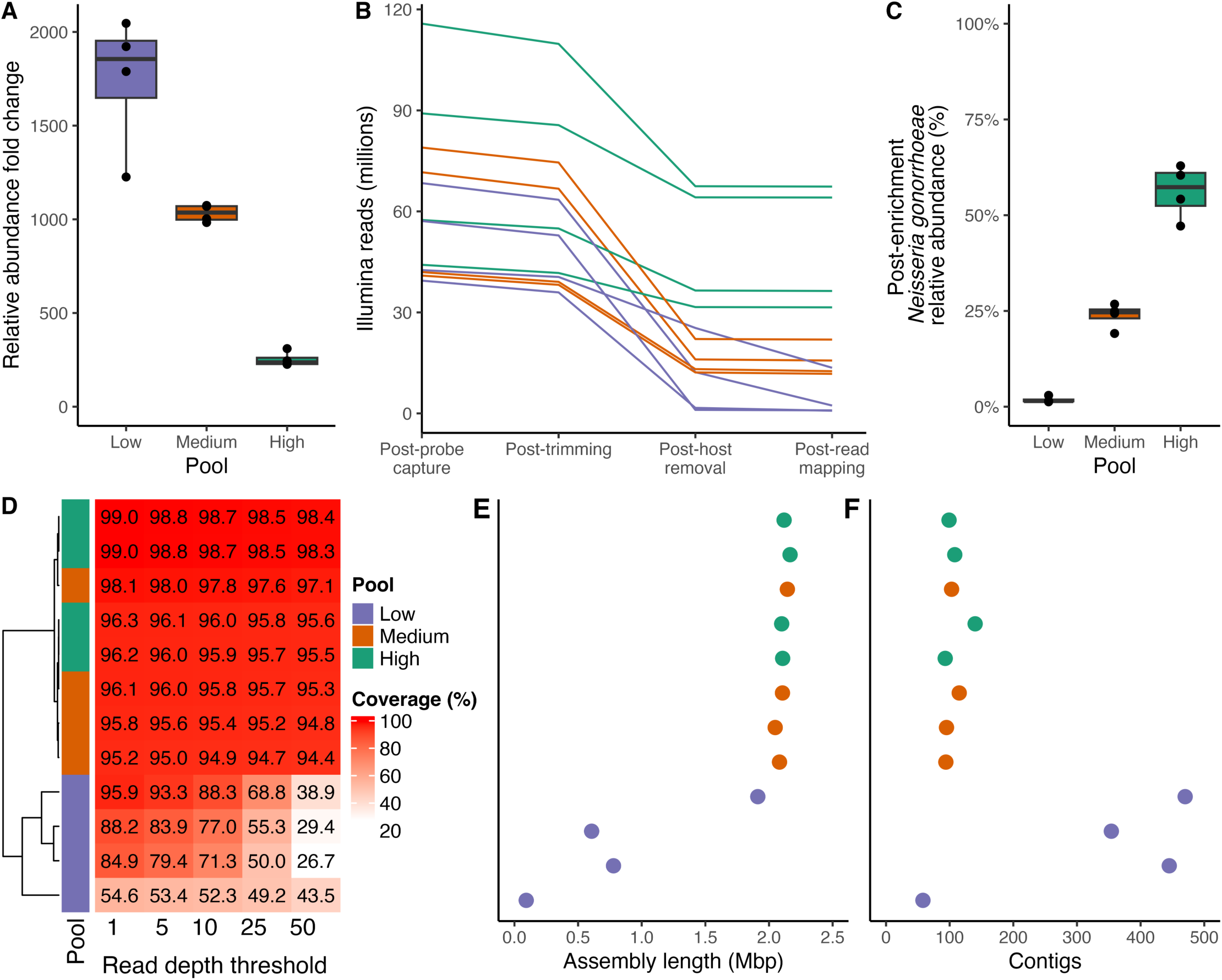
Application of probe-capture sequencing to clinical *Neisseria gonorrhoeae*-positive urine specimens. Probe-capture sequencing was performed on 12 *Neisseria gonorrhoeae*-positive urine cultures. (A) Fold change in pathogen abundance after probe capture, as determined by shallow sequencing using a portion of an Illumina NovaSeq X lane. (B) Read count progression through the bioinformatic pipeline, highlighting the impact of human read removal and pathogen-specific read retention. (C) Relative abundance of *N. gonorrhoeae*-assigned reads in the initial sample after probe-capture sequencing. (D) Heatmap detailing the proportion of reference genome coverage at various read depth thresholds. (E) Total genome assembly length of each sample. (F) Total number of contigs of each sample.

Alongside the recovery of genome assemblies, the molecular detection of sequence types and resistance-associated genotypes is critical to molecular surveillance of multidrug-resistant organisms. Inspection of the nine assemblies revealed genetic diversity among these isolates, with each assembly belonging to a distinct ST. Assembly-based screening using AMRsearch suggested successful detection of resistance determinants across major drug classes and mechanisms, including detection of conserved resistance-associated loci and specific alleles (**Figure 4A**). The median (IQR) contig size for resistance determinants was 43,077 bp (IQR, 54,287 bp), with only 23 / 164 (14.0%) being under 10 kbp (**Figure 4B**). The median (IQR) minimum distance of genotypes from contig barriers was 10,935.5 bp (IQR, 17,951.25 bp), with 17 / 164 (10.4%) being found under 1 kbp from a contig barrier (**Figure 4C**). Read mapping to Pathogenwatch reference genes revealed considerable chromosome-normalized non-zero coverage across most loci (**Figure 4D**) and at the specific positions where AMRSearch calls resistance mutations (**Figure 4E).** Breadth of coverage, defined as 90% of the loci at 20X, was detected for most genotype-containing assemblies (**Supplementary Figure 7)**. However, a few loci, notably the 16S rRNA gene, 23S rRNA gene, *bla_TEM_*, and *gyrA*, showed consistently poor chromosome-normalized median non-zero coverage across isolates, identifying them as priorities for future probe set optimization (**Supplementary Figure 8**). Collectively, probe-capture sequencing of clinical urine specimens can substantially improve pathogen read recovery, high-quality, short-read assemblies, and enable the detection of resistance determinants.

**Figure 4.**
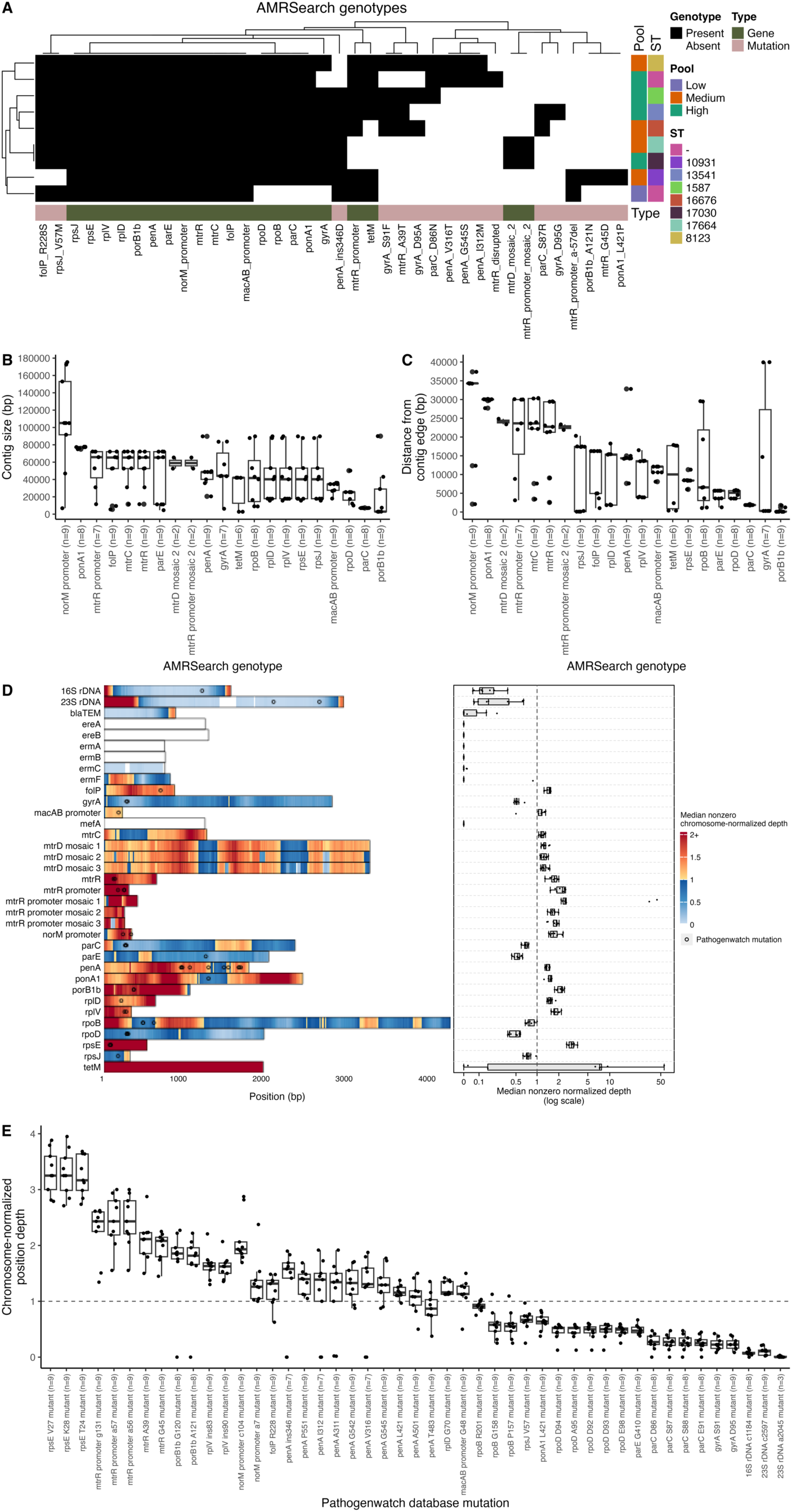
Detection of resistance-associated genotypes in *Neisseria gonorrhoeae*-positive urine specimens using probe-capture sequencing. Evaluation of probe-capture sequencing to detect resistance-associated genotypes in nine *Neisseria gonorrhoeae*-positive urine cultures with high-quality genome assemblies. (A) Assembly-based evaluation of resistance-associated genotype presence using Pathogenwatch’s AMRSearch. (B) Size of genotype-harboring contigs. (C) Minimum distance of genotypes from contig edges. (D) Read-based evaluation of resistance-associated genotypes was performed by read mapping to Pathogenwatch’s database. For each isolate, positional read depth was normalized by the median non-zero normalized depth detected on read mapping to the well-characterized *N. gonorrhoeae* FA 1090 reference genome. The heatmap shows the median value across isolates at each position, whereas the boxplots show the isolate-level normalized coverage for each locus. Circles denote mutation positions in the Pathogenwatch database, regardless of whether the mutation was detected. (E) Chromosome-normalized read depth at Pathogenwatch mutation positions across the nine isolates. Labels indicate the number of isolates with non-zero coverage at each mutation.

## DISCUSSION

Robust molecular epidemiology surveillance is required to accurately characterize the epidemiology and evolution of microbial pathogens. Challenging cultivation limits the feasibility of comprehensive, genomic surveillance of numerous pathogens with global public health burden, such as *N. gonorrhoeae*. In this study, we piloted the application of probe-capture sequencing to clinical urinary specimens positive for *N. gonorrhoeae*. Leveraging a probe set designed to capture the genomic diversity of publicly available genomes, this methodology substantially enriched pathogen-specific reads 200-to 2000-fold, enabling the recovery of high-quality genomic data across specimens with moderate- and high-levels of pathogen relative abundance in sample metagenomes. Enrichment was also observed for samples with low initial relative abundance of *N. gonorrhoeae*, though *de novo* assemblies of post-capture sequence data led to more fragmented genomes. These findings demonstrate the potential of probe-capture sequencing to characterize the burden and epidemiology of fastidious pathogens, with implications for strengthening the clinical and public health surveillance of this public health threat.

Molecular surveillance requires innovative sequencing approaches to overcome the limitations of conventional culture-dependent sequencing methods. While shotgun metagenome sequencing can be used to assemble genomes from multi-organism samples, the relative abundance of *N. gonorrhoeae* in urine samples was never sufficiently high to allow for genome assembly even with sequencing depths of 100M clusters. Thus, our application of probe-capture sequencing to clinical urinary specimens demonstrates that genomically and phenotypically representative probes can enable pathogen recovery across specimens with low and moderate pathogen abundance. Recent studies exemplify the breadth of probe-capture sequencing to inform clinical care and public health surveillance (42). Notably, probe-capture sequencing can increase sensitivity to detect antibiotic resistance genotypes and widen the diagnostic window for difficult-to-culture pathogens (9-12, 43). Alongside improvement of pathogen surveillance, probe-capture sequencing has been leveraged to investigate microbial evolution, ecology, and pathogenesis (13). Together, these applications highlight the growing potential of targeted enrichment as a flexible sequencing strategy to study many pathogens that remain difficult to characterize using conventional approaches.

Despite the promising performance of probe-capture sequencing to *N. gonorrhoeae*, several factors remain critical to investigate to establish the generalizability of this approach. First, the clinical specimens evaluated in this study were urinary specimens. Future studies should evaluate the performance of probe-capture sequencing across other anatomical sites, including extragenital sites (44). Other studies have reported positive enrichment on oropharyngeal positive specimens directly from urine and urethral specimens, albeit using fewer genome assemblies during probe-capture design (9, 45). Second, although enrichment improved recovery across most low-abundance specimens, one specimen did not yield sufficient on-target sequences for downstream assembly. Additional enrichment strategies, including sequential rounds of probe-capture, should be evaluated to improve genome recovery. Genomic diversity was restricted to the publicly available Pathogenwatch collections, which may hinder generalizability. Incorporation of newly sequenced genomes into probe designs could potentially further expand the breadth of sequence diversity captured in future iterations of probe sets. Third, the design of pathogen-specific probes reduced the ability to detect resistance-associated genotypes found across numerous species, such as 16S rRNA, 23S rRNA, and *bla*_TEM_ genes. Although this approach reduced potential capture of non-target genetic content, supplementation using other probe sets may be necessary to illuminate genetic content at these loci. Finally, we note that the design of hybridization capture probes required a quite large probe set to ensure broad coverage across a genetically diverse bacterial species. Further testing will examine higher-level multiplexing to reduce the cost/sample for this workflow.

In conclusion, our successful application of probe-capture sequencing to *N. gonorrhoeae* underscores the potential for targeted enrichment to inform the clinical management and surveillance of difficult-to-culture pathogens. The markedly increased pathogen recovery and successful genome assembly provide the foundation for expanding surveillance beyond conventional, culture-based sequencing methods. Future studies should evaluate the performance across diverse specimens to further generalize to other difficult-to-cultivate bacteria. Expanding this approach could further advance targeted enrichment as a powerful tool for studying pathogen diversity, transmission dynamics, and antibiotic resistance in public health surveillance.

## AUTHOR STATEMENTS

### Author contributions

S.J.G. and K.J.P. conceived and designed the study.

P.M.W., K.J.P., and S.J.G. facilitated the collection and processing of isolates and clinical data.

E.S.S. and K.J.P. supervised the research.

B.W.B. designed the probes.

K.J.G., A.S., D.B., and B.B. performed computational analyses.

K.J.G. and A.S. wrote the manuscript with input from all co-authors.

All authors read and approved the final manuscript.

### Ethical approval statement

The Institutional Review Board of the Rush University Medical Center approved this study with waiver of consent.

### Data availability

All data produced in the present study are available upon reasonable request to the authors.

## SUPPLEMENTARY MATERIALS

**Supplementary Figure 1.**
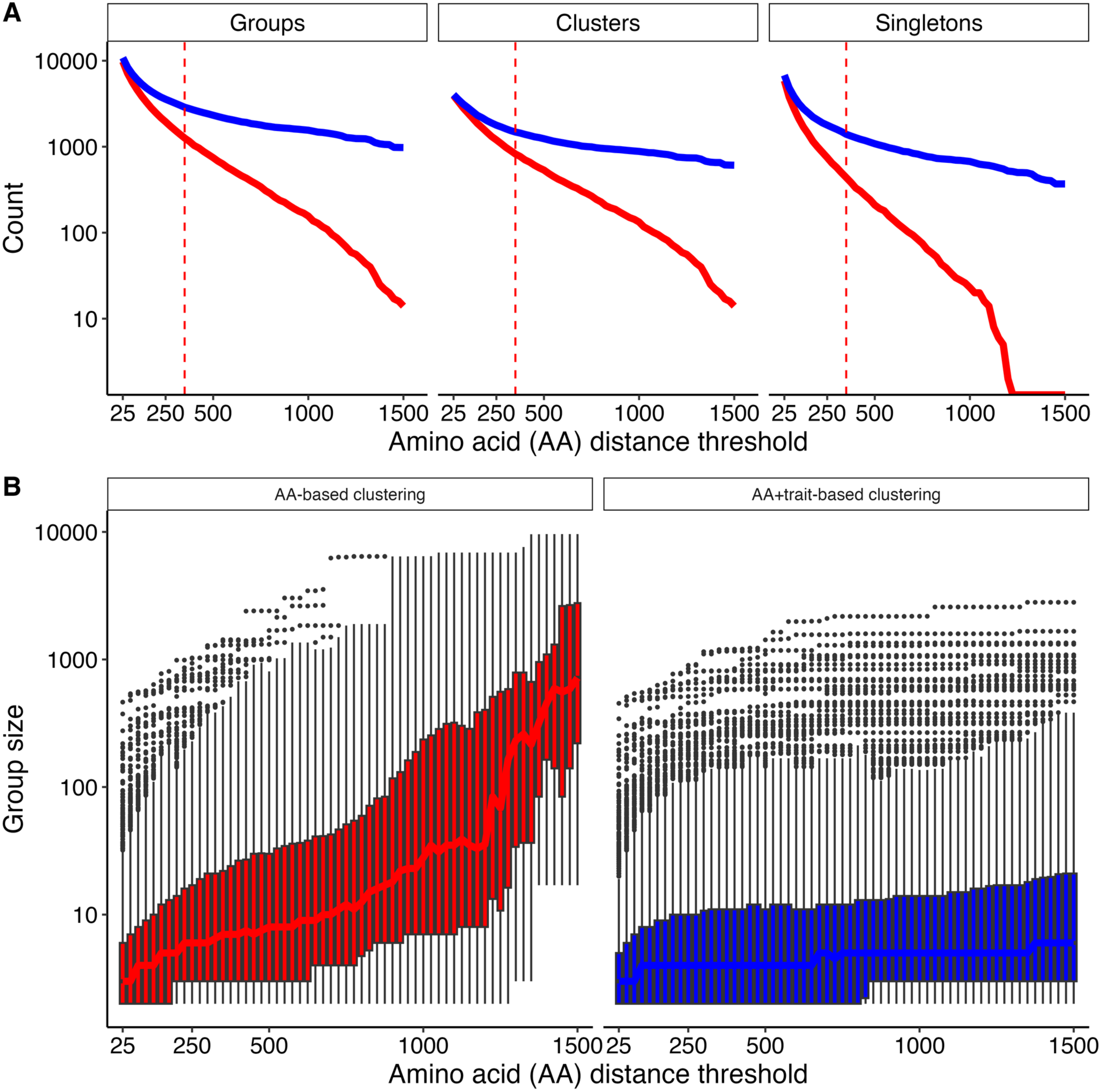
Influence of amino acid distance threshold choice on assembly clustering dynamics. Genome assemblies were clustered at various core gene amino acid (AA) distance thresholds, ranging from 25 to 1,500. After partitioning assemblies into genetically related clusters (AA-based clustering), multi-assembly clusters were further partitioned based on their resistance-associated genotype and phenotype patterns (AA+trait-based clustering). (A) The number of groups across distance thresholds and the two genome clustering steps. Clusters were defined as groups with more than one isolate. (B) Distribution of group sizes across distance thresholds depicted as a box-and-whisker plot and faceted by clustering step.

**Supplementary Figure 2.**
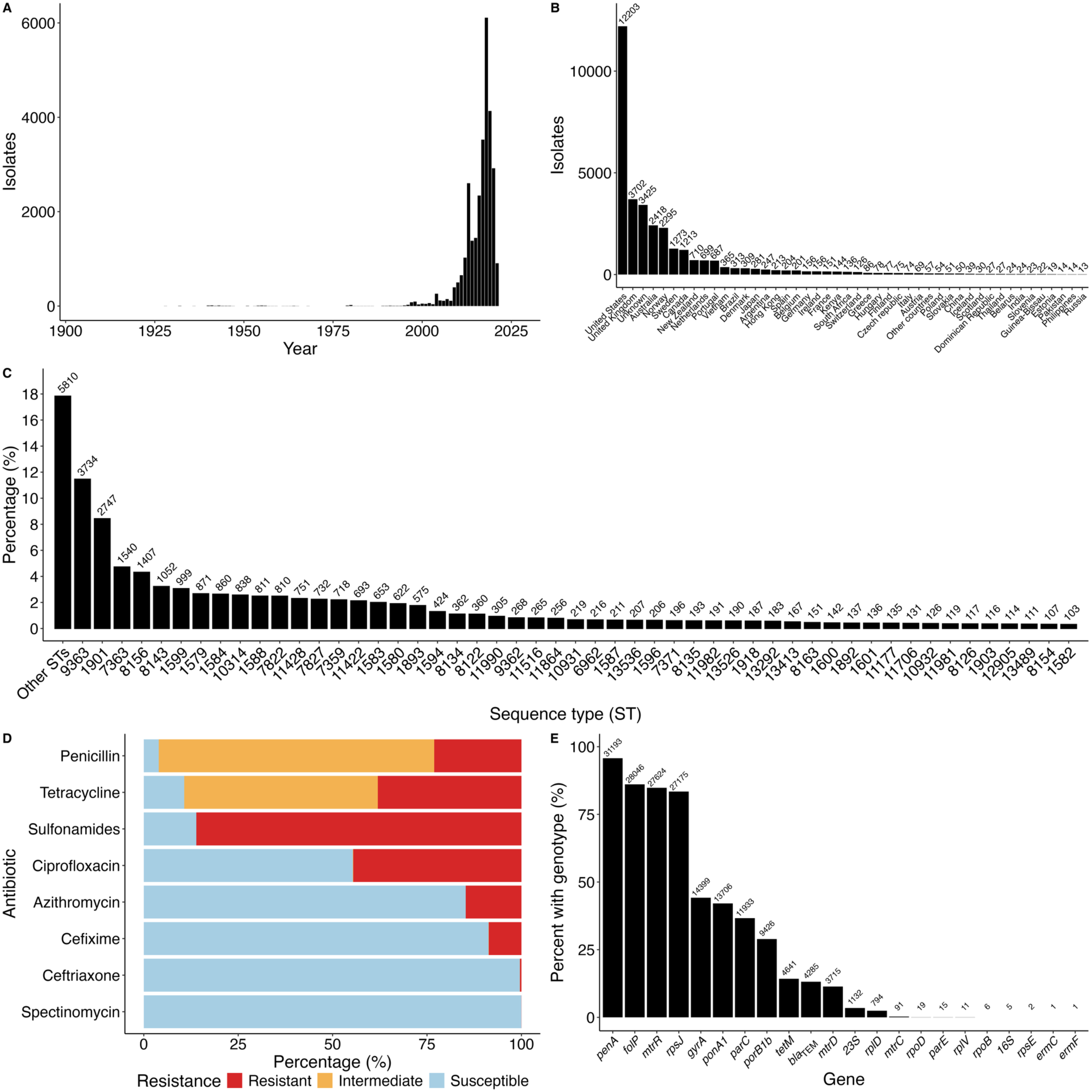
Overview of the Pathogenwatch *Neisseria gonorrhoeae* isolate collection. The global isolate collection comprises all publicly available whole-genome sequences available on Pathogenwatch at the time of analysis (downloaded on 11/24/2024). (A) Histogram of collection dates for selected genome assemblies. (B) Distribution of genome assemblies across countries. Countries with under ten assemblies were classified as ‘Other countries’ to aid visualization. (C) Proportion of assemblies by sequence type (STs). To aid in visualization, only the top 51 STs are represented. (D) Antibiotic susceptibility profiles of selected assemblies, categorized as resistant, intermediate, or susceptible, as predicted by Pathogenwatch. (E) Proportion of assemblies with resistance-associated genotypes, collapsed at the locus level.

**Supplementary Figure 3.**
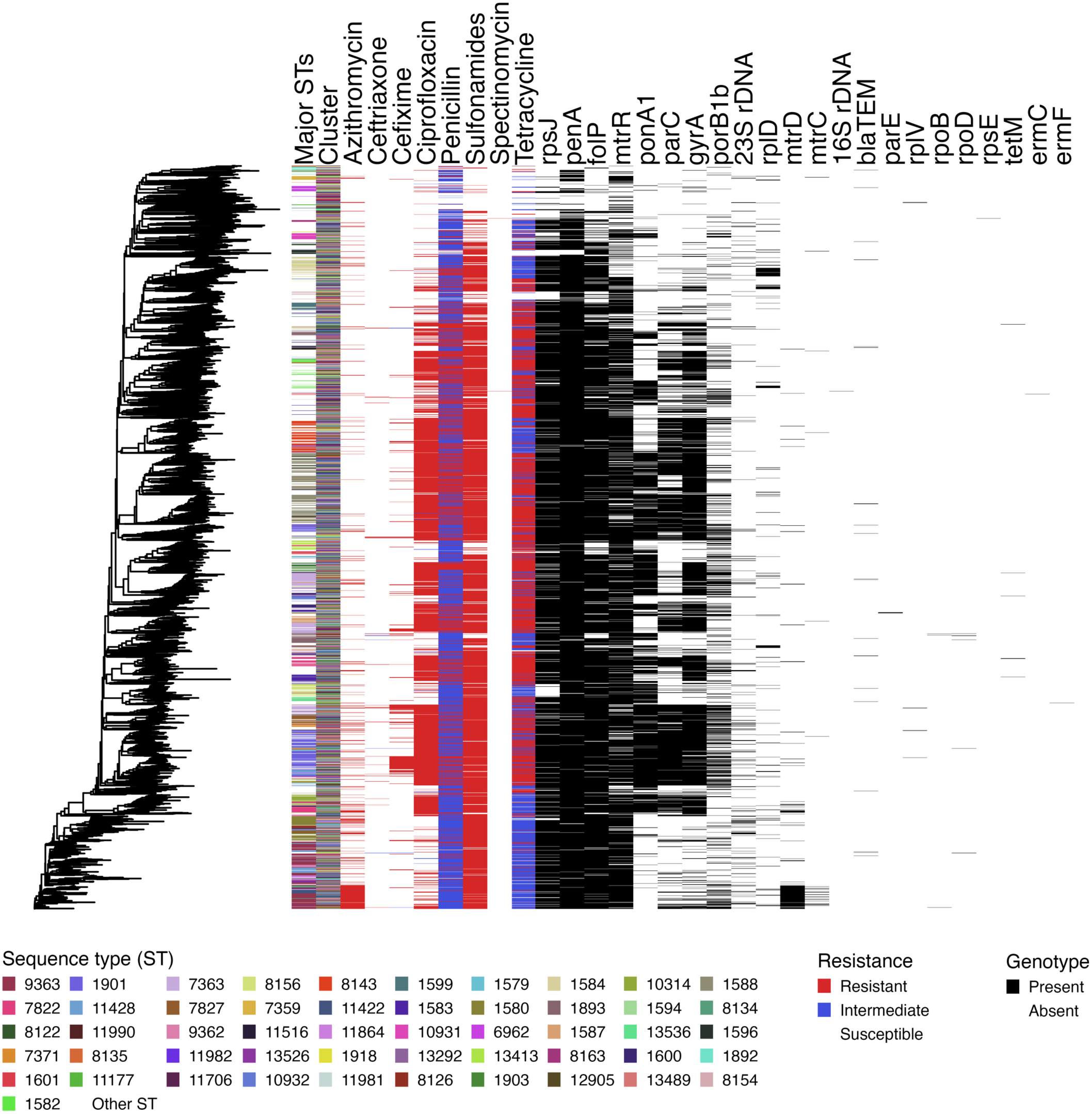
Phylogeny of the representative assemblies chosen for probe set generation. Phylogenetic visualization of the Pathogenwatch *Neisseria gonorrhoeae* assemblies used for probe set generation. First, the top 51 sequence types (STs), determined by their frequency in the overall collection, were displayed. Next, the amino acid distance clusters, inferred using a threshold distance of 350, were overlaid on the phylogeny. Next, antibiotic susceptibility profiles were overlaid for eight antibiotics, classified as susceptible, intermediate, or resistant, as predicted by Pathogenwatch. Finally, the presence of resistance-associated genotypes, aggregated at the locus level, was overlaid on the phylogeny.

**Supplementary Figure 4.**
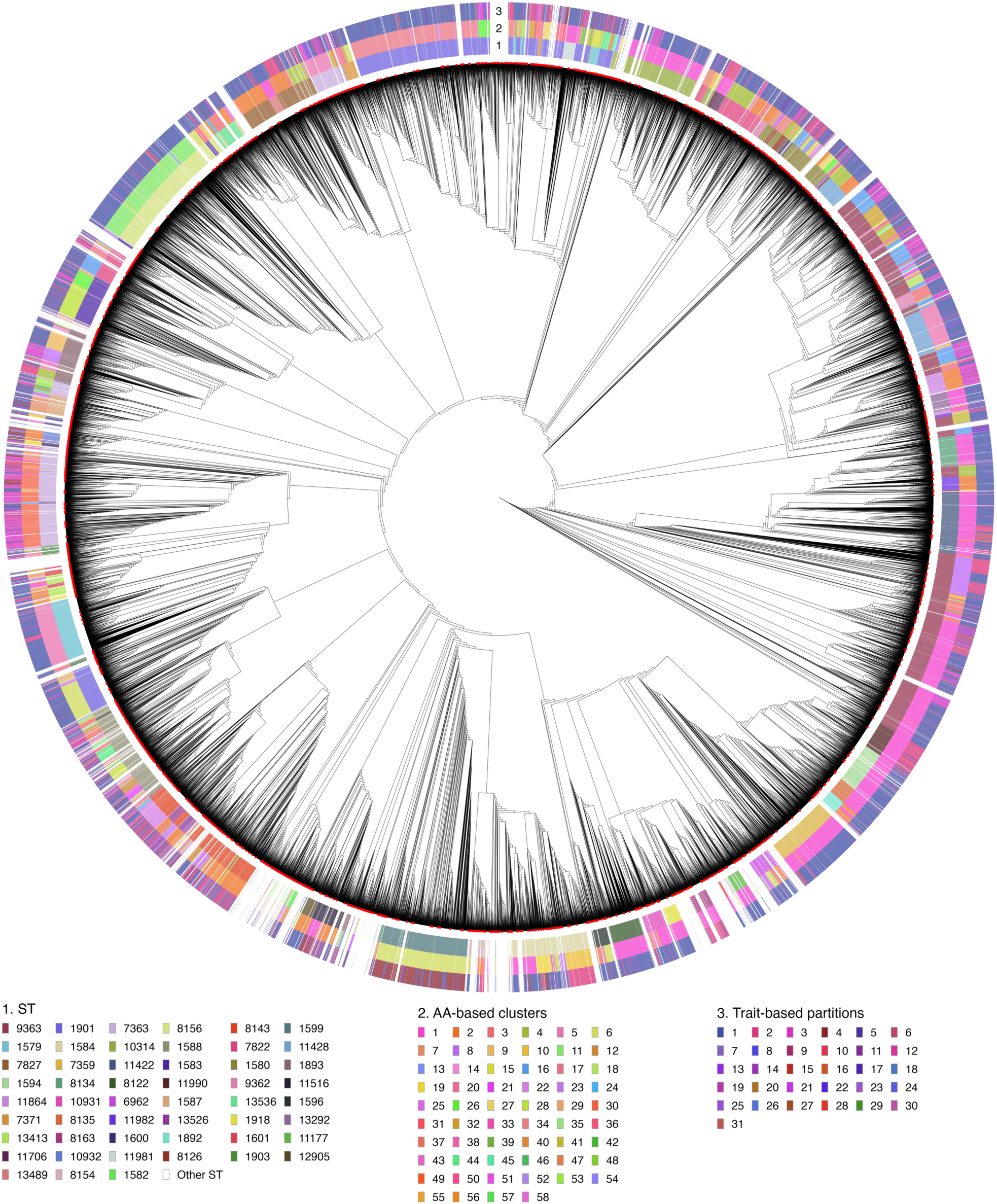
Amino acid– and trait-based partitioning of *Neisseria gonorrhoeae* genomes. Phylogeny of publicly available whole-genome sequences available on Pathogenwatch at the time of analysis (downloaded on 11/24/2024). Amino acid distance-based clustering and subsequent partitioning of clusters by resistance-associated genotypes and phenotype predictions were performed to group clusters into representative groups. To facilitate representation of the groups, amino acid distance- and trait-based partitions are sequentially numbered within each sequence type. To simplify data visualization, partitioning data from the top 51 most abundant sequence types are provided.

**Supplementary Figure 5.**
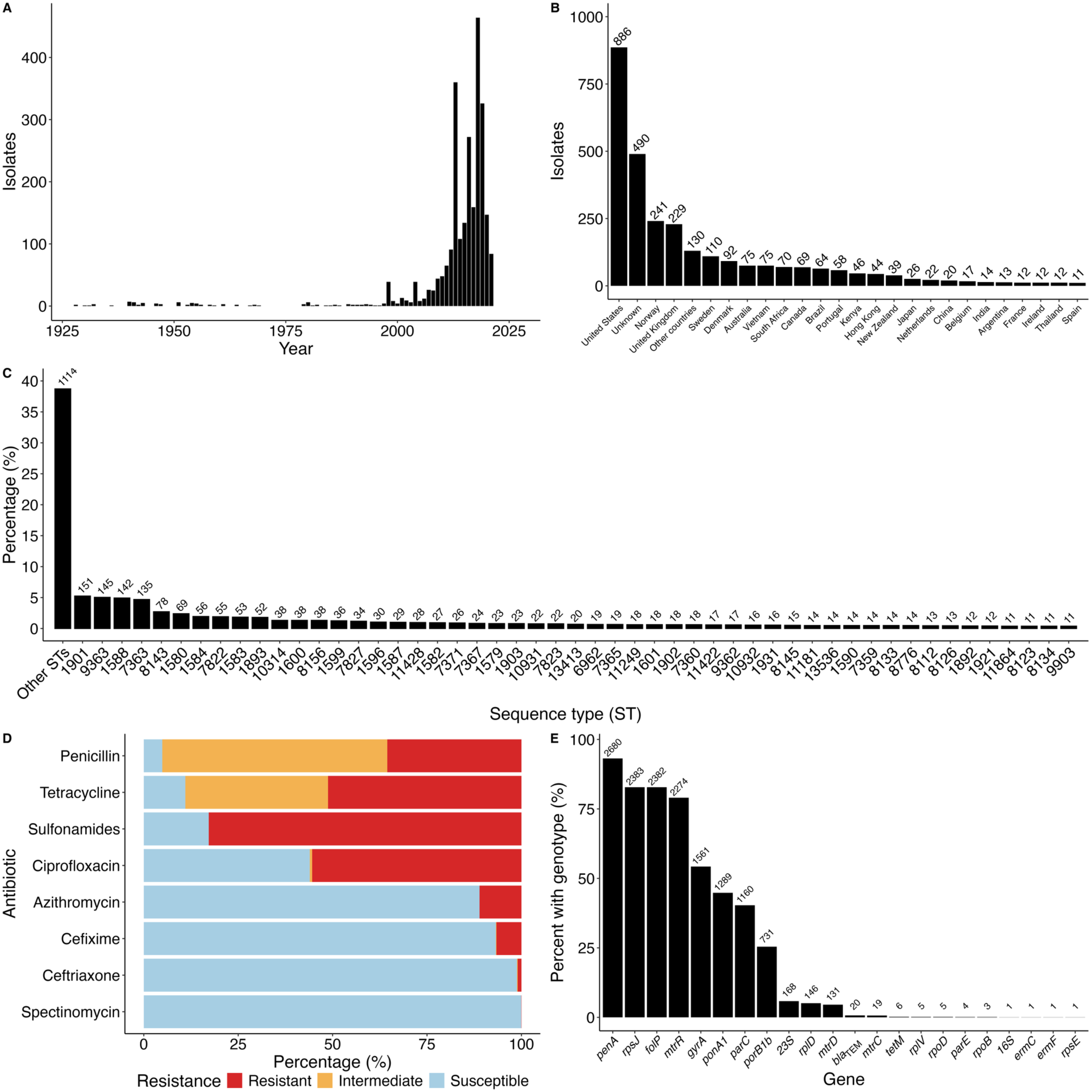
Overview of the *Neisseria gonorrhoeae* genomes selected for probe set generation. Representative isolates from Pathogenwatch, downloaded on 11/24/2024, were chosen using amino acid-based clustering and subsequent trait-based partitioning of cluster isolates. (A) Histogram of collection dates for selected genome assemblies. (B) Distribution of genome assemblies across countries. Countries with under ten assemblies were classified as ‘Other countries’ to aid visualization. (C) Proportion of assemblies by sequence type (STs). To aid in visualization, only the top 51 STs are represented. (D) Antibiotic susceptibility profiles of selected assemblies, categorized as resistant, intermediate, or susceptible, as inferred by Pathogenwatch. (E) Proportion of assemblies with resistance-associated genotypes, collapsed at the locus level.

**Supplementary Figure 6.**
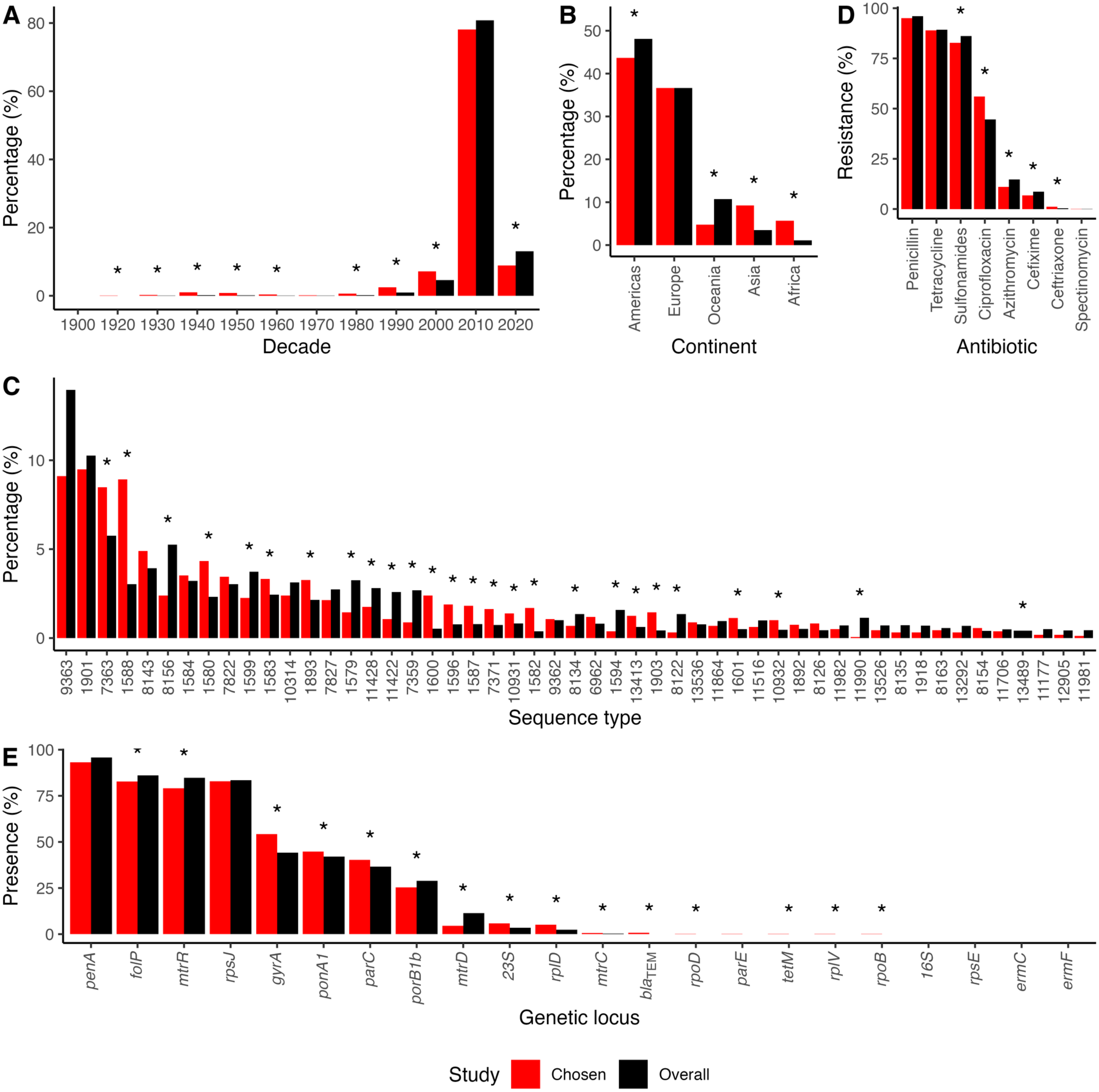
Representative genomes compared to the full *Neisseria gonorrhoeae* collection. To evaluate the impact of partitioning assemblies using genetic distance and resistance-associated patterns, we compared the representative collection used for probe design to the overall Pathogenwatch collection: (A) temporal distribution of isolates by decade, (B) geographic distribution by continent, (C) sequence type (ST) distributions, (D) antibiotic susceptibility profiles, and (E) the distribution of resistance-associated loci at the gene level. Resistance was defined as a phenotypic prediction in the intermediate or resistant category. Asterisks indicate statistically significant differences between the chosen and global collection (Fisher’s exact test, p-value <0.05).

**Supplementary Figure 7.**
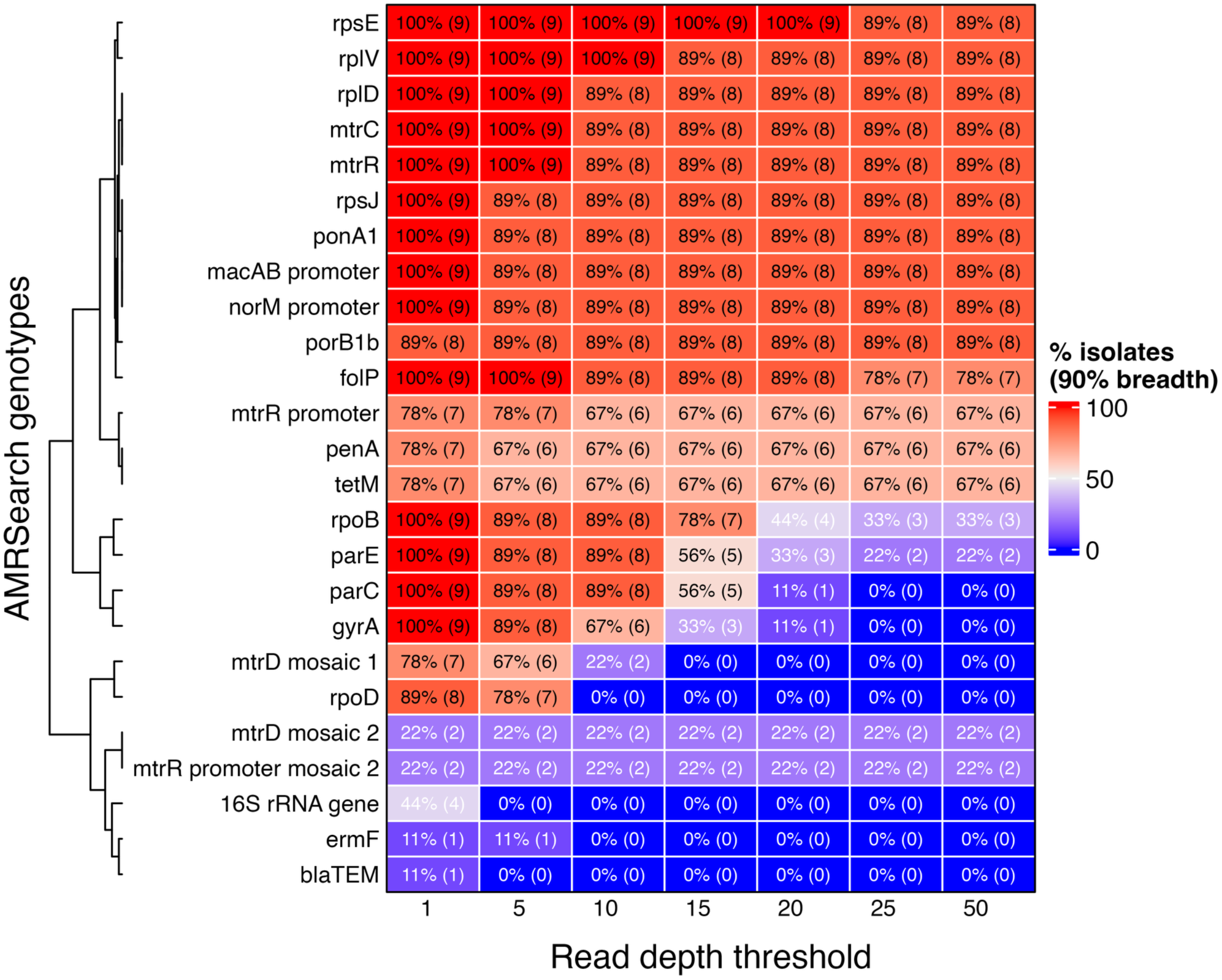
Coverage across resistance-associated reference loci at various read depth thresholds. Read mapping was performed to evaluate coverage of Pathogenwatch’s AMRsearch resistance-associated loci among nine *Neisseria gonorrhoeae*-positive urine cultures with high-quality assemblies. For each locus with at least one genome with non-zero coverage, the proportion of bases with read depth at or above the specified threshold was calculated. A locus was considered adequately covered if at least 90% met the depth threshold.

**Supplementary Figure 8.**
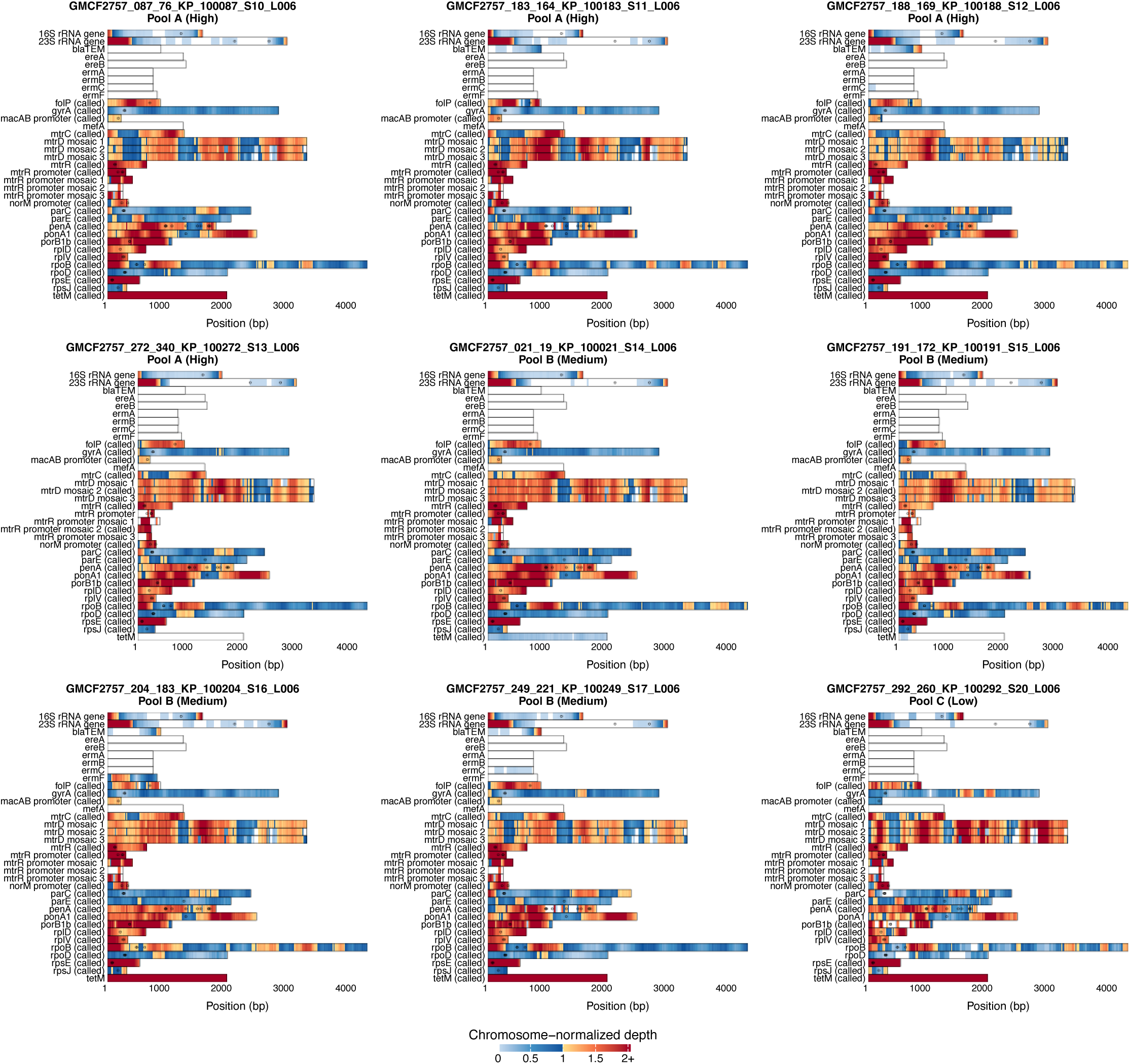
Chromosome-normalized coverage across resistance-associated reference loci. Read mapping was performed to evaluate coverage patterns of Pathogenwatch resistance-associated reference loci across nine *Neisseria gonorrhoeae*-positive urine cultures with high-quality genome assemblies. Each panel represents one isolate, with read depth at each position normalized using the median non-zero coverage when mapping reads to the well-characterized *N. gonorrhoeae* FA 1090 reference genome. Loci detected by AMRSearch are labeled “(called)”, while circles denote mutation positions in the Pathogenwatch database, regardless of mutation detection.

